# Study of the lymphocyte change between COVID-19 and non-COVID-19 pneumonia cases suggesting other factors besides uncontrolled inflammation contributed to multi-organ injury

**DOI:** 10.1101/2020.02.19.20024885

**Authors:** Yishan Zheng, Zhen Huang, Guoping Yin, Xia Zhang, Wei Ye, Zhiliang Hu, Chunmei Hu, Hongxia Wei, Yi Zeng, Yun Chi, Cong Cheng, Feishen Lin, Hu Lu, Lingyan Xiao, Yan Song, Chunming Wang, Yongxiang Yi, Lei Dong

## Abstract

**Background:** The corona virus disease 2019 (COVID-19) shows unusually high transmission rate and unique clinical characteristics, with key pathological mechanism remaining unclear. Here, we analysed the laboratory data based on clinical samples from COVID-19 patients, in parallel comparison with non-COVID-19 pneumonia cases, in an attempt to elucidate the key pathological features of COVID-19 during its infection of the human body.

**Methods:** We analysed biochemical indices and lymphocyte subpopulation in COVID-19 patients, and compare these data from non-COVID-19 pneumonia cases. Correlation analysis was performed between leukocyte subgroups count and biochemical indexes in COVID-19 patients.

**Results:** The study enrolled 110 patients, comprising 88 COVID-19 patients and 22 non-COVID-19 pneumonia cases. We observed significant differences, including abnormal biochemical indices (CRP, LDH, AST, eGFR, and sodium ion concentration) and reduced lymphocyte subsets count, between the COVID-19 patients and non-COVID-19-caused pneumonia cases. Correlation analysis indicates that the count for lymphocyte subsets-but not that for neutrophils and monocytes-exhibits a significant negative correlation with biochemical indices relating to organ injury, in the COVID-19 infected patients.

**Conclusions:** The study indicates significantly different clinical features between 2019 novel coronavirus (2019-nCoV)-caused and non-2019-nCoV-caused pneumonia, especially in terms of lymphocytopenia and organ injury. Notably, correlation analysis demonstrates that tissue damage in COVID-19 patients is attributed to virus infection itself rather than uncontrolled inflammatory responses (“cytokine storm”). These findings provide new insights for developing efficient therapeutic strategies against COVID-19 infection.

## Introduction

Since its initial outbreak in the Chinese city Wuhan in early December 2019, the corona virus disease 2019 (COVID-19) has spread throughout China and globally, with the total number of reported cases approaching 100,000.^1^ Analysis of clinical data available is in urgent demand for revealing the mechanisms of tissue damage and further devising more specific and efficient treatment strategies.

The 2019 novel coronavirus (2019-nCoV) is a novel beta coronavirus based on the data from gene sequencing.^2^ Compared with other beta coronaviruses, such as SARS-CoV and MERS-CoV, 2019-nCoV infection leads to significant differences in clinical symptoms, according to the latest studies ^[3]^, including remarkably higher occurrence of non-fever patients, fewer upper respiratory and gastrointestinal symptoms, a different profile of increased blood cytokines, more frequent multi-organ injury, and lower death rate.^3-5^ These differences, along with its unusual high transmission rate, suggest a different pathological mechanism underlying its infection of the human body.

In this study, we analyzed the clinical and laboratory data of most of the 2019-nCoV-infected patients in the China city Nanjing, aimed at revealing the unique pathological features of COVID-19 cases in comparison with non-2019-nCoV-caused pneumonia cases. Importantly, through correlation analysis of the changes in blood cell subgroups and key blood biochemical indexes of tissue injury, we have discovered unexpected clues of the pathogenic process following 2019-nCoV infection, which may provide valuable information for designing efficient approaches of treatment.

## Methods

### Patients

A study based on 88 COVID-19 cases and 22 non-COVID-19 pneumonia cases from the Second Hospital of Nanjing (Tangshan Branch, 1 Kangfu Road) was performed. All COVID-19 cases were diagnosed based on the World Health Organization (WHO) interim guidance and the Diagnosis and Treatment Plan for Novel Coronavirus Pneumonia Cases (Provisional) (7th Ed.) ^6-7^ and confirmed positive for 2019-nCoV nucleic acid in the respiratory samples via real-time reverse-transcriptase polymerase-chain-reaction (RT-PCR). All non-COVID-19 pneumonia cases samples were negative, based on the absence of 2019-nCoV nucleic acid in the respiratory tract specimen. The RT-PCR assay was performed according to the protocol established by the WHO.^8^ Based on pulmonary CT imaging findings, all patients with 2019-nCoV infection were graded 1-3 from mild to server according to the lung injury degree. This study was approved by the ethics committee of the Second Hospital of Nanjing, with written informed consent obtained from each patient.

### Blood samples and laboratory data collection

Blood samples from patients were collected for laboratory assessments and flow cytometry according to the doctor’s instruction. All patients didn’t receive any treatment before blood sampling. Laboratory data were obtained from electronic medical records. Laboratory assessments comprised the following indexes: complete blood count, blood biochemical test, liver, and renal function, electrolytes, C-reactive protein (CRP), procalcitonin, lactate dehydrogenase, and creatine kinase.

### Flow cytometry analysis

Peripheral blood mononuclear cells (PBMC) were collected from EDTA anticoagulant whole blood via erythrocyte lysis. PBMCs were incubated with a panels of fluorescence-labeled antibodies (CD45/CD3/CD4/CD8, CD45/CD3/CD16CD56/CD19), or corresponding IgG controls (Biolegend, San Diego, CA, USA) and rinsed with 1*PBS. Finally, the samples were examined by a flow cytometry analyzer (BD FACSCalibur, BD Biosciences, San Jose, CA, USA).

### Statistical analysis

The results are expressed as means ± SD. Data were statistically analyzed using Prism SPSS 19.0 (SPSS Inc., Chicago, IL, USA) and assessed for normality or homogeneity of variance. For normally distributed continuous variables, differences between multiple groups were compared using one-way ANOVA with Dunnett’s tests or, if appropriate, using one-way ANOVA with post-hoc Bonferroni correction. Continuous variables that were not normally distributed were compared using Kruskal-Wallis test. Spearman correlation analysis between the blood cell subgroups and the blood biochemical indexes were conducted using SPSS 19.0. A value of p < 0.05 was considered significant. NS = not significant.

## Results

### Laboratory findings of COVID-19 infected patients

A total of 110 patients were included in the study (Table 1), consisting of 88 2019-nCoV infected patients (47 males and 41 females) with a mean age of 46.386±17.532 and 22 non-COVID-19 pneumonia cases (11 males and 11 females) with a mean age of 36.727±9.948. The cohort of 2019-nCoV infected cases were further divided into the following three groups from mild to severe accroding to CT imaging findings: grade 1 group (aged 39.444±14.328, 6 males and 3 females), grade 2 group (aged 44.845±16.799, 36 males and 35 females) and grade 3 group (aged 67.875±12.229, 5 males and 3 females).

**Table 1.** Clinical characteristics and laboratory findings of patients with 2019-nCoV infection (n=88) and non-COVID-19 pneumonia patients (n=22).

| Parameter (characteristics laboratory findings) | 2019-nCoV infected patients (case groups) |  |  | Non-COVID-19 pneumonia patients<br>n=22 |
| --- | --- | --- | --- | --- |
|  | Grade 1<br>n=9 | Grade 2<br>n=71 | Grade 3<br>n=8 |  |
| Patients No. | 9 | 71 | 8 | 22 |
| Age (years) | 39.444±14.328 | 44.845±16.799 | 67.875±12.229* | 36.727±9.948 |
| Gender: male/female | 6/3 | 36/35 | 5/3 | 11/11 |
| Blood leukocyte count, 10<sup>9</sup>/L | 5.028±2.52 | 4.772±1.406 | 4.525±1.596 | 6.006±1.837 |
| Lymphocyte count, 10<sup>9</sup>/L | 1.678±0.817 | 1.379±0.55* | 1.22±0.807* | 1.927±0.441 |
| Lymphocyte ratio, % | 33.6±6.511 | 30.149±10.504 | 28.063±15.759 | 33.532±10.24 |
| Neutrophil count, 10<sup>9</sup>/L | 2.939±1.639 | 2.905±1.25 | 2.465±0.91 | 3.779±1.869 |
| Neutrophil ratio, % | 57.889±8.04 | 60.134±11.369 | 63.875±17.697 | 57.436±11.668 |
| Monocyte count, 10<sup>9</sup>/L | 0.366±0.163 | 0.383±0.123 | 0.3±0.111 | 0.434±0.158 |
| Monocyte ratio, % | 7.667±2.415 | 8.544±2.954 | 8.063±3.478 | 7.355±2.579 |
| Platelet count, 10<sup>9</sup>/L | 224.889±48.766 | 191.718±55.575* | 152.375±43.471* | 242.045±64.865 |
| Mean platelet volume, fL | 10.144±0.972 | 10.352±1.361* | 10.313±0.81 | 9.1±1.133 |
| Erythrocyte sedimentation rate, mm/h | 17.75±12.881 | 16.681±15.412 | 33±23.523 | 11.571±15.397 |
| Hematocrit, mm/h | 39.489±5.215 | 40.679±5.338 | 40.05±4.534 | 41±5.408 |
| C-reactive protein level, mg/L | 4.218±4.671 | 5.612±4.005 | 10.798±5.566* | 3.762±4.105 |
| Procalcitonin, ng/ml | 0.053±0.096 | 0.096±0.495 | 0.07±0.057 | 0.046±0.052 |
| Album level, g/L | 46.267±1.862 | 44.282±3.862 | 40.3±5.279 | 45.959±5.333 |
| Globulin level, g/L | 21.856±1.914 | 21.468±3.514 | 21.15±3.215 | 22.759±3.868 |
| Total bilirubin, µmol/L | 11.067±4.808 | 12.323±4.754 | 13.875±5.14 | 14.936±5.741 |
| Ischemia Modified Albumin(IMA), U/mL | 72.889±4.256 | 74.269±14.21 | 74.875±14.545 | 79.773±15.964 |
| Lactose dehydrogenase, U/L | 233±89.285 | 228.634±78.901 | 295.375±78.425* | 199±50.143 |
| Creatinine kinase, U/L | 104.111±96.883 | 105.537±128.146 | 74.875±65.392 | 102.409±135.612 |
| **Alanine aminotransferase, U/L** | 30.033±24.279 | 27.025±24.145 | 24.188±12.676 | 34.559±25.066 |
| **Aspartate aminotransferase, U/L** | 25.989±15.554 | 24.694±12.76 | 33.813±13.547 | 23.05±8.904 |
| **Lactic acid level, mmol/L** | 2.912±0.918 | 2.302±1.065 | 1.825±0.902 | 3.12±1.369 |
| **Estimated glomerular filtration rate, ml/min** | 117.63±15.011 | 116.062±16.958 | 96.953±12.143* | 122.898±13.615 |
| **Potassium, mmol/L** | 3.876±0.443 | 3.877±0.387 | 3.855±0.363 | 4.015±0.374 |
| **Sodium, mmol/L** | 139.667±2.458 | 139.142±3.058 | 136.525±2.187* | 140.509±2.732 |

Plus-minus values are means ± SD.
\* P<0.05, P-value stands for the comparison between 2019-nCoV infected patients (Grade 1, Grade 2, or Grade 3) and non-COVID-19 patients.

The complete blood count results indicated that the number of lymphocytes and platelets were significantly decreased in both grade 2- and grade 3-group COVID-19 patients, in comparison with non-2019-nCoV-infected pneumonia patients, suggesting that COVID-19 patients could have lymphopenia and thrombocytopenia. In contrast, no significant differences in the quantities and proportions of monocytes and neutrophils were observed among these groups. Grade 3-group COVID-19 patients had significantly higher levels of C-reactive protein (CRP), compared with non-2019-nCoV-infected pneumonia patients. Blood biochemistry detection found that several indices related to organ injuries, including lactose dehydrogenase (LDH), aspartate aminotransferase (AST), estimated glomerular filtration rate (eGFR), and sodium ion concentration, were significantly different between grade 3-group COVID-19 patients and non2019-nCoV-infected pneumonia patients.

### Circulating lymphocyte subset profile of COVID-19 patients

To further explore the mechanism, and since lymphopenia was observed in COVID-19 patients, we performed flow cytometry to analyze the composition of lymphocyte subpopulations in the peripheral blood from patients. As shown in Table 2, the count of total CD45+ lymphocytes, CD3+ lymphocytes, CD4+ T cells, CD8+ T cells, and CD19+ B cells in both grade 2- and grade 3-group COVID-19 patients were significantly lower than those in non-COVID-19-infected pneumonia patients. However, the differences of the proportions of CD3+/CD45, CD4+/CD45, CD8+/CD45, CD4+/CD8+, and CD16CD56+/CD45+ and CD4+CD8+ T cells count were only observed between grade 3-group COVID-19 patients and non-2019-nCoV-infected pneumonia patients. Meanwhile, no difference of CD16CD56+ NK cells count was observed among different groups.

**Table 2.** Results of lymphocyte subpopulations in peripheral blood of patients with 2019-nCoV infection (n=88) and non-COVID-19 pneumonia patients (n=22).

| Parameter (characteristics<br>laboratory findings) | 2019-nCoV infected<br>patients (case groups) |  |  | Non-COVID-19 pneumonia patients<br>n=22 |
| --- | --- | --- | --- | --- |
|  | Grade 1 | Grade 2 | Grade 3 |  |
|  | n=9 | n=71 | n=8 |  |
| CD45+ lymphocytes count,<br>cells/uL | 1958.778±887.919 | 1563.704±631.897* | 1186.25±767.487* | 2169.091±522.489 |
| CD3+ T cells count, cells/μL | 1433.333±803.57 | 1117.563±465.484* | 530.25±255.417* | 1605.545±426.538 |
| CD3+/CD45+ ratio, % | 71.159±12.3 | 71.415±9.774 | 49.864±15.247* | 73.765±7.834 |
| CD4+ T cells count, cells/μL | 695.333±420.085 | 613.31±276.869* | 336.375±140.798* | 918.182±275.854 |
| CD4+/CD45+ ratio, % | 35.556±9.888 | 39.211±9.58 | 29.875±9.79* | 42.045±6.869 |
| CD8+ T cells count, cells/μL | 567.778±377.568 | 416.62±213.984* | 161.125±109.694* | 589.227±200.706 |
| CD8+/CD45+ ratio, % | 27.434±9.314 | 26.617±8.218 | 14.306±7.998* | 27.182±6.551 |
| CD4+/CD8+ (Th/Ts ) ratio, % | 1.421±0.589 | 1.666±0.773 | 3.164±2.277* | 1.653±0.537 |
| CD4+CD8+ T cells count,<br>cells/μL | 1.889±1.9 | 2.099±1.742 | 0.375±0.744* | 2.818±2.737 |
| CD4+CD8+/CD45 ratio, % | 0.094±0.093 | 0.137±0.123 | 0.029±0.054 | 0.123±0.118 |
| CD19+ B cells count, cells/μL | 227.444±142.755 | 200.225±211.713* | 109.5±57.756* | 326.273±134.208 |
| CD19+ B cells ratio, % | 11.782±3.838 | 12.051±5.785 | 11.448±5.666 | 14.779±4.326 |
| CD16+CD56+ NK cells count,<br>cells/μL | 288±206.573 | 229.93±168.416 | 537.5±674.557 | 221.909±144.364 |
| CD16+CD56+ NK cells ratio, % | 16.533±12.726 | 15.47±10.782 | 38.2±18.427* | 10.774±6.624 |

Plus-minus values are means ± SD.
\* P<0.05, P-value stands for the comparison between 2019-nCoV infected patients (Grade 1, Grade 2, or Grade 3) and non-COVID-19 patients.

### Correlation analysis between leukocyte subgroups count and biochemical indexes

To examine the role of inflammatory responses in organ injury of COVID-19 patients, we performed Spearman’s rank correlation coefficient (ρ) analysis between leukocyte subgroups count and biochemical indexes relating to organ injury in both COVID-19 (Table 3) and non-2019-nCoV-infected pneumonia patients (Table 4). In COVID-19 patients, LDH, AST, and sodium ion concentration showed significant correlations (P<0.05) with total lymphocyte and all lymphocyte subsets count; meanwhile, creatinine kinase (CK), total bilirubin, eGFR, and potassium ion concentration only exhibited significant correlations with part of lymphocyte subpopulation count (Table 3). Among the lymphocyte subsets, CD4+ Th cells count showed significant correlations with most biochemical indices (Fig 1). In contrast, no significant correlation was found between biochemical indexes and the counts for monocytes and neutrophils. Meanwhile, LDH only negatively correlated with CD3+ T cell count and CD8+ T cell count and AST exhibited with CD8+ T cell count in non-2019-nCoV-infected pneumonia patients (Table 4). In general, correlation analysis implied a significant negative correlation between lymphocyte subsets count and organ injury degree in COVID-19 patients rather than non-2019-nCoV-infected pneumonia patients.

**Table 3.** Spearman correlation analysis between biochemical index levels and the count of lymphocyte subpopulations, neutrophil and monocyte in patients with 2019-nCoV infection (n=88).

| Biochemical indexes | CD45+ lymphocytes count |  | CD3+ T cells count |  | CD4+ T cells count |  | CD8+ T cells count |  | CD19+ B cells count |  | Neutrophils count |  | Monocytes count |  |
| --- | --- | --- | --- | --- | --- | --- | --- | --- | --- | --- | --- | --- | --- | --- |
| | $\rho$ | P value | $\rho$ | P value | $\rho$ | P value | $\rho$ | P value | $\rho$ | P value | $\rho$ | P value | $\rho$ | P value |
| Lactose dehydrogenase | -0.297 | 0.005 | -0.387 | <0.001 | -0.437 | <0.001 | -0.251 | 0.018 | -0.322 | 0.002 | 0.159 | NS | -0.059 | NS |
| Creatinine kinase | -0.166 | NS | -0.198 | NS | -0.243 | 0.022 | -0.126 | NS | -0.075 | NS | 0.111 | NS | -0.020 | NS |
| Alanine aminotransferase | -0.121 | NS | -0.161 | NS | -0.162 | NS | -0.086 | NS | -0.081 | NS | 0.098 | NS | 0.038 | NS |
| Aspartate aminotransferase | -0.320 | 0.002 | -0.388 | <0.001 | -0.420 | <0.001 | -0.288 | 0.006 | -0.337 | 0.001 | 0.046 | NS | -0.086 | NS |
| Total bilirubin | -0.061 | NS | -0.073 | NS | -0.047 | NS | -0.110 | NS | -0.214 | 0.046 | -0.080 | NS | 0.133 | NS |
| Estimated glomerular filtration rate | 0.191 | NS | 0.252 | 0.018 | 0.256 | 0.016 | 0.249 | 0.019 | 0.178 | 0.098 | -0.144 | NS | 0.021 | NS |
| Sodium | 0.276 | 0.009 | 0.370 | <0.001 | 0.436 | <0.001 | 0.243 | 0.023 | 0.407 | <0.001 | 0.017 | NS | 0.046 | NS |
| Potassium | 0.353 | <0.001 | 0.321 | 0.002 | 0.175 | NS | 0.328 | 0.002 | 0.185 | NS | 0.099 | NS | 0.099 | NS |

$\rho$ = Spearman's rank correlation coefficient. NS = non-significant.

**Table 4.** Spearman correlation analysis between biochemical index levels and the count of lymphocyte subpopulations, neutrophil and monocyte in non-COVID-19 pneumonia patients (n=22).

| Biochemical indexes | CD45+ lymphocytes count |  | CD3+ T cells count |  | CD4+ T cells count |  | CD8+ T cells count |  | CD19+ B cells count |  | Neutrophils count |  | Monocytes count |  |
| --- | --- | --- | --- | --- | --- | --- | --- | --- | --- | --- | --- | --- | --- | --- |
| | $\rho$ | P value | $\rho$ | P value | $\rho$ | P value | $\rho$ | P value | $\rho$ | P value | $\rho$ | P value | $\rho$ | P value |
| Lactose dehydrogenase | -0.312 | NS | -0.506 | 0.016 | -0.416 | NS | -0.489 | 0.021 | -0.167 | NS | 0.049 | NS | 0.094 | NS |
| Creatinine kinase | 0.123 | NS | 0.079 | NS | -0.163 | NS | 0.184 | NS | 0.002 | NS | 0.057 | NS | 0.371 | NS |
| Alanine aminotransferase | 0.007 | NS | -0.185 | NS | -0.231 | NS | -0.070 | NS | 0.038 | NS | -0.037 | NS | 0.190 | NS |
| Aspartate aminotransferase | -0.262 | NS | -0.419 | NS | -0.464 | 0.030 | -0.244 | NS | -0.244 | NS | -0.244 | NS | 0.126 | NS |
| Total bilirubin | 0.347 | NS | 0.306 | NS | 0.238 | NS | 0.414 | NS | 0.297 | NS | 0.362 | NS | 0.109 | NS |
| Estimated glomerular filtration rate | 0.342 | NS | 0.359 | NS | 0.321 | NS | 0.318 | NS | 0.057 | 0.098 | -0.028 | NS | -0.213 | NS |
| Sodium | 0.190 | NS | 0.198 | NS | 0.073 | NS | 0.232 | NS | 0.090 | NS | 0.205 | NS | 0.272 | NS |
| Potassium | 0.171 | NS | 0.166 | NS | 0.335 | NS | 0.197 | NS | 0.114 | NS | -0.041 | NS | -0.156 | NS |

$\rho$ = Spearman's rank correlation coefficient. NS = non-significant.

**Figure 1.**
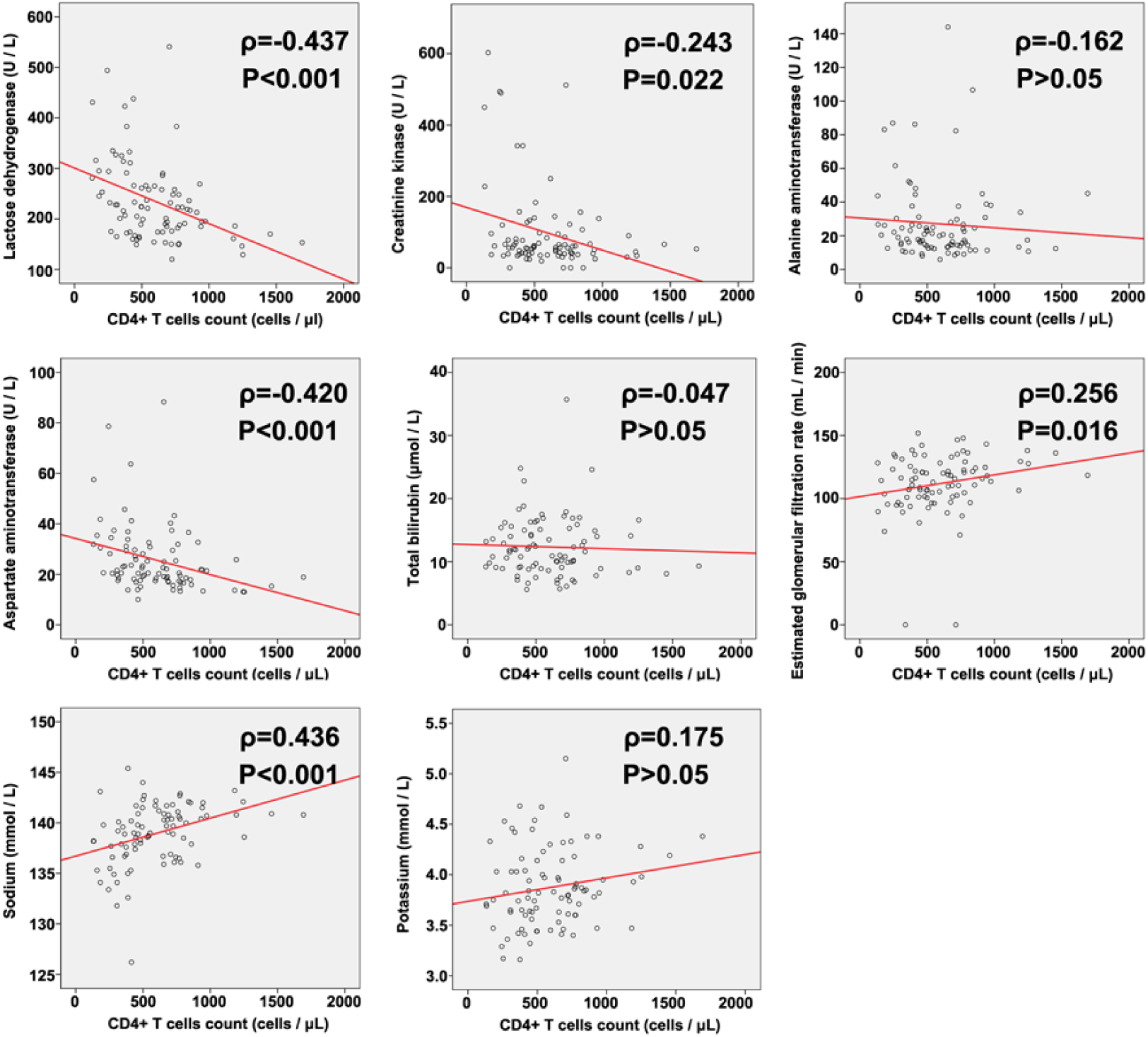
Spearman’s rank correlation coefficient (ρ) analysis between CD4+ T cells count and biochemical indexes in patients with 2019-nCoV infection. A value of p < 0.05 was considered statistically significant.

## Discussion

Our most striking finding is that lymphocytopenia, which is common in acute infections,^9^ is particularly significant in 2019-nCoV infections.^3^ Others have demonstrated the correlation between the lymphocytopenia and the clinical severity.^10^ Most previous reports only compared the severe groups against non-severe patients or heath controls; to further study this clinical feature, in our study, we compared the differences in the subgroups of lymphocytes harvested from COVID-19 patients with those from pneumonia patients not infected by 2019-nCoV. We noticed that lymphocytes also decreased in the grade 2-group COVID-19 patients, suggesting that it might not only be a consequence of the infection but is probably a critical factor driving the development and deterioration of the disease. Among the lymphocyte subgroups, B cell exhibited the most significant differences from the non-2019-nCoV-infected pneumonia patients. Since B cells are responsible for the humoral immunity against the invaders via producing antibodies,^11^ the impotent response of B cells may fail to restrict the virus expansion and release of free virions *in vivo*, which promotes the infection.

Another new and notable feature discovered is that the change in the proportion of the CD45+ lymphocytes was insignificant, compared with non-COVID-19 patients. Recently, postmortem results also found the numbers of trilineage in the central immune system (bone marrow) and lymphocytes in the peripheral immune system (spleen and lymph node) were all decreased significantly, indicating the peripheral lymphocytopenia cannot be simply attributed to the change of lymphocyte tissue distribution.^7^ Thses findings, first, demonstrate that the infection impacts on different lymphocyte subgroups evenly, and more interestingly, suggest that abnormality in cells that regulate the adaptive immunity may be responsible for the reduction of lymphocytes. For instance, H5N1 infection causes lymphocytopenia through dendritic cell (DC) dysfunction,^12^ suggesting the function of key antigen-presenting cells can be drastically impaired during the infection. According to a study in rats, the myeloid cells also express the 2019-nCoV receptor ACE2;^13^ thus, we should consider the possibility of the virus infecting these cells, which is supported by the significant decrease in the platelet counts in both grade 2-group and grade 3-group COVID-19 patients and the reduced count of trilineage in the bone marrow.

Moreover, recent imaging findings and postmortem biopsies reports indicate that the infection can potentially damage other organs than the lung, including the kidney, heart, liver and the testis.^14-16^ In correspondence with pathological findings, we also notice changes in multiple biochemical indexes relating to organ injuries, including LDH, CK, AST, and eGFR. In SARS-Cov cases, the multi-organ injury was considered as a result of the inflammatory reaction (inflammatory cytokine storm). However, the early report about COVID-19 cases found that the profile of cytokine in the blood of patients was different from that from SARS-Cov cases: not only pro-inflammatory cytokines but also anti-inflammatory cytokines (IL-4, IL-10, etc.) increased in the COVID-19 cases.^4^ Additionally, all recent reports demonstrated that high fever cases (>38°C) were significantly fewer (even in severe cases) in COVID-19 patients than in SARS-CoV or MERS-CoV cases.^3^ These features suggest that the consequences in the COVID-19 patients might not be mainly due to the inflammatory reaction, especially in the non-severe cases. Instead, the damage was likely to be caused by the virus itself. Therefore, we performed a correlation analysis between the number of lymphocyte subgroups and the biochemical indexes. We found that most of the indexes relating to organ injuries were negatively correlated with the number of lymphocytes in 2019-nCoV infected patients but not in non-2019-nCoV-infected pneumonia patients, further highlighting the virus infection – rather than the inflammatory reaction – as the possible cause of the multi-organ injury. Another supportive finding is that the difference in the number of monocytes, neutrophils, and NK cells is not significant in our analysis; while theoretically, these innate immune cells are the main player in a “cytokine storm”. Interestingly, the most significant correlation happened between the injury indexes and CD4+ Th cell counts. Because CD4+ Th cells are critical for the regulation of both cellular immunity and humoral immunity,^17^ it is reasonable that these cells are most sensitive to the total anti-virus immune responses. This finding further suggests that the damage of the tissue is from the virus itself. Consistently, studies on the tissue distribution of ACE2 suggest that the virus receptor is widely expressed in human tissue including the digestive tract, kidney, testis, and others.^18^

We highlight that the above finding is important for the design of treatment regimens. Current treatments do not suggest using specific drugs to improve the activity of immune systems, such as the GM-CSF or other immune stimuli, in fearing of the exaggeration of inflammatory reactions and cytokine storm.^19^ However, based on our clinical findings, we should seriously consider using immune-activating treatments for COVID-19 patients, which will be helpful to compensate the dysfunctions in the adaptive immune system and accelerate virus clearance process *in vivo*. Considering the high possibility of secondary infection induced by excessive inflammation in the severe cases, this strategy should be safer to apply in non-severe patients with the aim to lower the occurrence of severe cases.

Taken together, our analysis in the present study, based on clinical data, demonstrate significant differences between pneumonia caused and not caused by 2019-nCoV-especially including the lymphocytopenia. Our results further suggest that the tissue damage to COVID-19 patients might be mainly due to virus infection itself rather than uncontrolled inflammatory reactions. These findings are valuable for the development of more efficient treatments.

## Data Availability

The data used to support the findings of this study are available from the corresponding author upon request.

## Acknowledgments

This study was funded by the Young Medical Talents of Jiangsu Province (QNRC2016056), the fifth period “333 high-level personnel training project” in Jiangsu Provience (2016-III-0084), the National Natural Science Foundation of China (31971309 and 31671031), the Jiangsu Province Funds for Distinguished Young Scientists (BK20170015).

